# Spatial and Temporal Hotspot Analysis of COVID-19 in Toronto

**DOI:** 10.1101/2024.08.30.24312852

**Authors:** Afia Amoako, Mabel Carabali, Erjia Ge, Ashleigh R Tuite, David N Fisman

**Affiliations:** Division of Epidemiology, Dalla Lana School of Public Health, University of Toronto, 155 College St Room 500, Toronto, Ontario M5T 3M7 Canada; Department of Epidemiology, Biostatistics, and Occupational Health, McGill University, 1020 Pine Avenue West, Montreal, Quebec, H3A 1A2, Canada; Centre for Immunization Programs, Public Health Agency of Canada, 130 Colonnade Rd A.L. 6501H Ottawa, Ontario, K1A 0K9, Canada

**Keywords:** COVID-19, Social Determinants of Health, GIS, Disease Mapping, Spatial Epidemiology

## Abstract

The COVID-19 pandemic in Toronto, Canada was unequal for its 2.7 million residents. As a dynamic pandemic, COVID-19 trends might have also varied over space and time. We conducted a spatiotemporal hotspot analysis of COVID-19 over the first four major waves of COVID-19 using three different applications of Moran’s I to highlight the variable experience of COVID-19 infections in Toronto, while describing the potential impact of socioeconomic and sociodemographic factors on increased risk of COVID-19 exposure and infection. Results highlight potential clustering of COVID-19 case rate hot spots in areas with higher concentrations of immigrant and low-income residents and cold spots in areas with more affluent and non-immigrant residents during the first three waves. By the fourth wave, case rate clustering patterns were more dynamic. In all, a better understanding of the unequal COVID-19 pandemic experience in Toronto needs to also consider the dynamic nature of the pandemic.

**HIGHLIGHTS:**

- The COVID-19 pandemic was spatially and temporarily dynamic in the City of Toronto.
- At first, hotspots were concentrated in areas with more marginalized residents.
- Later, COVID-19 spatial trends diverged from initially identified patterns.
- East Asian enclaves in the city disproportionally had lower COVID-19 case counts.
- COVID-19 studies need to consider the dynamic nature of the pandemic.

## INTRODUCTION

The COVID-19 experience in Toronto, Canada’s most populous city, has been varied and dynamic for its 2.7 million residents.(Statistics Canada, 2023) Early in the pandemic, it was evident that COVID-19 cases and outcomes were disproportionally concentrated amongst the socially and economically marginalized residents of the city. (Allen, 2020; Cheung, 2020; City of Toronto, 2020; Ingen et al., 2022; McKenzie et al., 2021) Such observations prompted an increased interest in the social, economic, and demographic disparities in COVID-19 cases in Ontario with the hopes of gaining a better understanding of the pandemic and focusing attention on communities with the highest risk. (Abdi et al., 2021; McKenzie, 2020) Spatial methods provide detailed visualization of infectious diseases and their distribution within a certain geographic location with tools that allow us to explore relationships between infection and potential risk factors, which can help inform community-level interventions.(Caprarelli and Fletcher, 2014; Lin and Wen, 2022) Within the context of Toronto, various studies have confirmed the spatial disparities of COVID-19 cases and outcomes across the city with a higher concentration of cases in locales with more racialized, immigrant, lower income and less educated residents. (Forsyth et al., 2023; Ma et al., 2022; Mishra et al., 2022; Nazia et al., 2022; Vaz, 2021; Wang et al., 2023; Xia et al., 2022)

However, the few spatiotemporal studies highlighting the disparities in the spatial distribution of COVID-19 have either focused on earlier months of the pandemic or taken a more cumulative approach over space and time. (Forsyth et al., 2023; Mishra et al., 2022; Nazia et al., 2022) The first two years of the SARS-CoV-2 pandemic were characterized by multiple distinct waves (Figure 1) due in part to new and often more infectious SARS-CoV-2 variants, the implementation of varying public health measures for each wave, and the varying availability and uptake of vaccines. (McCoy et al., 2020; Tsou et al., 2023; Vernon-Wilson et al., 2023)

**Figure 1:**
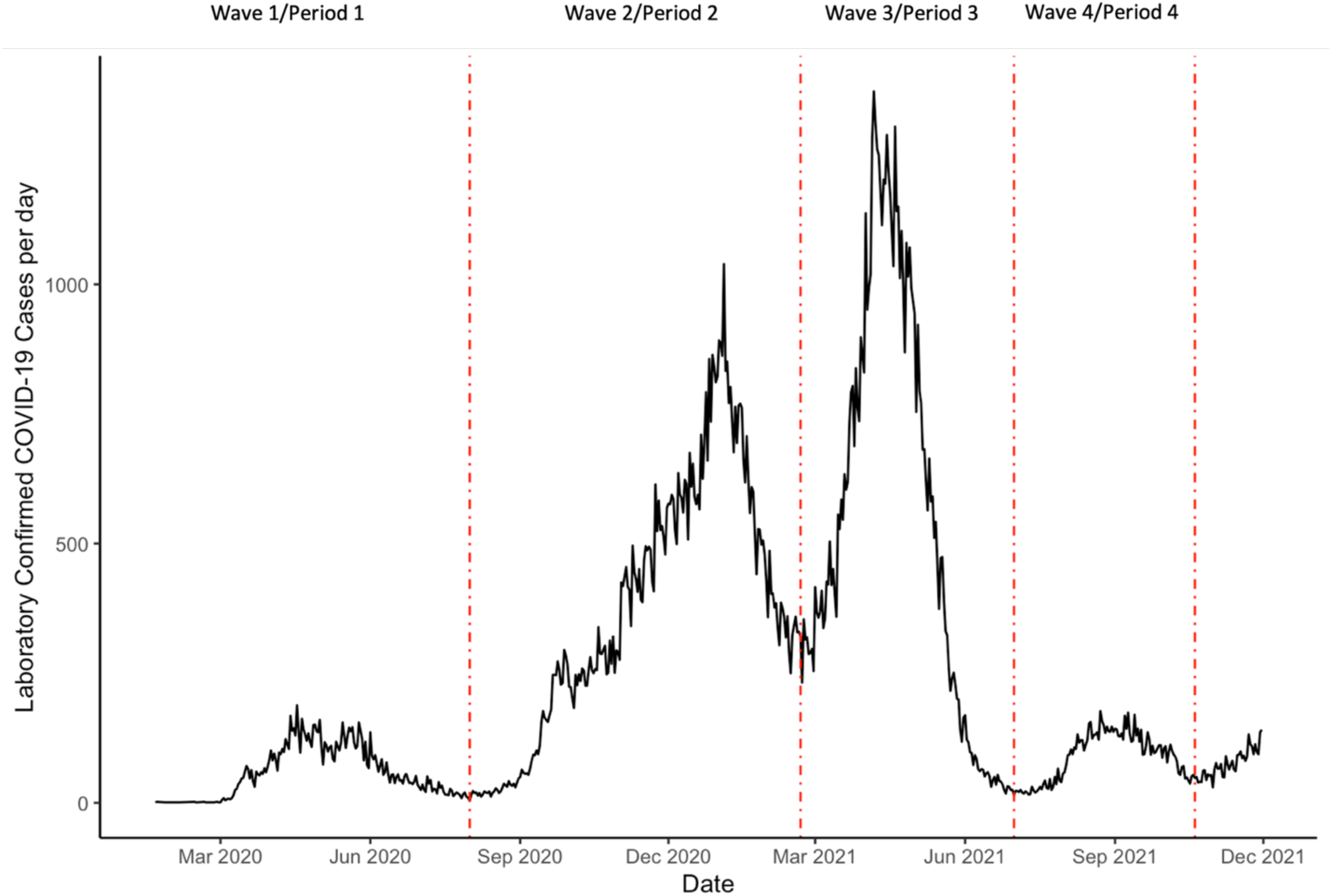
Daily Laboratory Confirmed COVID-19 cases in Toronto from January 2020 - December 2021. Waves of COVID-19 are indicated based on the patterns of peaks and valleys in case counts.

Our study hopes to contribute to the growing number of spatial studies that have explored the relationship between socioeconomic and sociodemographic factors and the spatial disparities of COVID-19 spatial Toronto. We performed a spatiotemporal descriptive hot spot analysis of COVID-19 case rates in Toronto with an emphasis on the dynamic experience of COVID-19 in the city. We intended to answer two pertinent questions: was the spatial distribution of COVID-19 cases consistent or variable over time given the dynamic experience of the pandemic? Were social demographic factors a potential indicator of what the spatial distribution of COVID-19 cases looked like over time?

## MATERIALS AND METHODS

### Study Population

The City of Toronto is Canada’s largest city with a diverse and multicultural population. More than half of the population identifies as an immigrant to Canada and/or as a visible minority. (City of Toronto, 2022a) Our analysis focused on residents of the City of Toronto who did not live in long-term care (LTC) facilities during our study period. Long-term care residents were excluded from analysis as the COVID-19 epidemic unfolded differently in these locations, with factors such as facility size, and over-representation of an at-risk population in one location, impacting infection transmission. (Konetzka et al., 2021) A general description of the population in Toronto can be found in Table 1. Our geographical unit of interest was census tracts primarily because they are geographical areas that consist of 2,500 – 8,000 people with similar socioeconomic characteristics. (Government of Canada, 2021)

**Table 1:** Descriptive Statistics of the City of Toronto in 2021 detailing general sociodemographic and socioeconomic characteristics of city residents along with average COVID-19 case rates per COVID-19 wave.

| **Table 1: Descriptive Statistics of the City of Toronto** (City of Toronto, 2022a, 2022c; Statistics Canada, 2023) |  |
| --- | --- |
| Total population in Toronto (2021) | 2,794,356 |
| Median Population per census tract (2021) | 4,666 |
| Age (2021) |  |
| Median Age in Toronto (years) | 39.6 |
| Proportion of Population over 65 years | 0.17 |
| Sociodemographic/Socioeconomic Characteristics (2021) |  |
| Median Income per Individual (Canadian Dollars) | 39,200 |
| Proportion of population that identify as visible minority | 0.557 |
| Proportion of population that identify as immigrants | 0.466 |
| Proportion of the population aged 25-64 with post-secondary education | 0.276 |
| Proportion of households in Toronto that are multigenerational | 0.035 |
| Average COVID case rate per 100,000 people (excluding LTC) |  |
| First Wave (January 20, 2020 – July 31, 2020) | 394.51 |
| Second Wave (August 1, 2020 – February 19, 2021) | 2666.3 |
| Third Wave (February 20, 2021 – June 30, 2021) | 2510.3 |
| Fourth Wave (August 2021 – October 20, 2021) | 358.38 |

### Study Design

We conducted an ecological study to assess the spatial and temporal distribution of COVID-19 case rates (per 100,000) at the census tract level from January 20, 2020, to October 20, 2021. Time was assessed in two ways. First, we divided our overall study period into four time periods to reflect the first four complete major waves of COVID-19 in Canada (See Figure 1 and Table 2) (Public Health Ontario, 2021, p. 2) Given these temporal units of analysis, each census tract had four COVID-19 case rates, one for each wave. We used these four waves to assess the presence of spatial autocorrelation as well as local hot spot analysis.

**Table 2:**
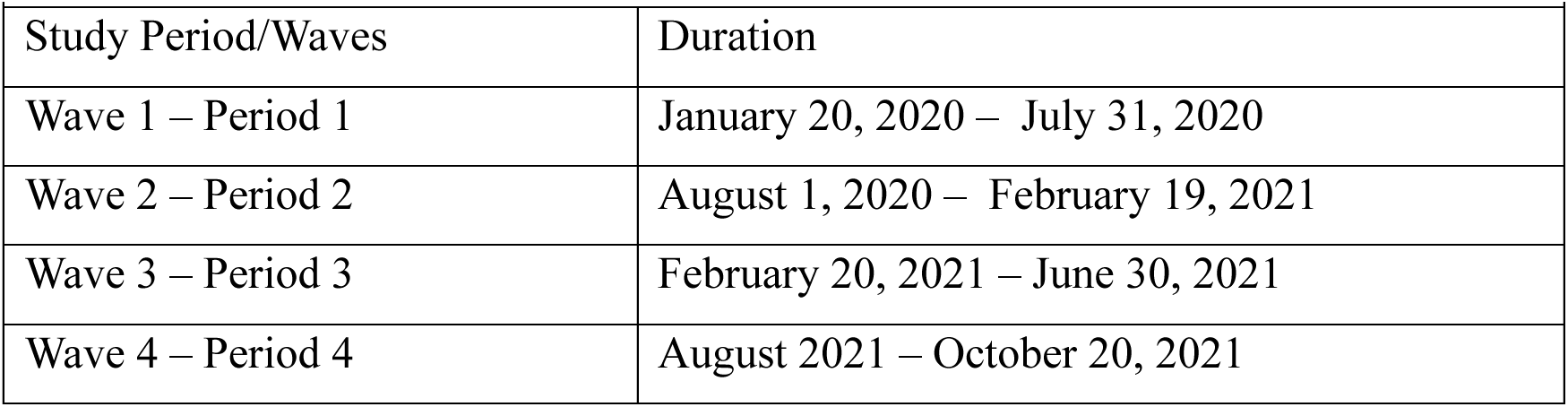
Duration of each of the first four major COVID-19 waves in Toronto.

| **Table 2: COVID-19 waves from January 2020 – October 2021** |  |
| --- | --- |
| Study Period/Waves | Duration |
| Wave 1 – Period 1 | January 20, 2020 – July 31, 2020 |
| Wave 2 – Period 2 | August 1, 2020 – February 19, 2021 |
| Wave 3 – Period 3 | February 20, 2021 – June 30, 2021 |
| Wave 4 – Period 4 | August 2021 – October 20, 2021 |

**Table 3:** Local Moran’s I indicate four different clustering patterns based on COVID-19 case concentration rates per census tract. Hotspots are identified as pink, cold spots are identified as light blue, while outliers are bright red for high-low outliers and bright blue for low-high outliers. Census tracts with no statistically significant clustering patterns are white.

| **Table 3: Local Moran's I Cluster Pattern** <sup>1-3</sup> |  |  |
| --- | --- | --- |
| Clustering Pattern |  | Definition |
|  | Hot Spot (high-high) | Census tracts that had similarly high COVID-19 case rates like their neighbours |
|  | Cold Spot (low-low) | Census tracts that had similarly low concentrations of COVID-19 case rates like their neighbours |
|  | High - Low Outlier | Census tracts with a higher concentration of COVID-19 case rates but surrounded by census tracts with lower COVID-19 case rates concentration |
|  | Low – High Outlier | Census tract with a lower concentration of COVID-19 case rates but surrounded by census tracts with higher COVID-19 case rate concentration |

For a cumulative understanding of the spatial and temporal distribution of COVID-19 case rates in the city over our study period, we divided our overall study period into 4-week intervals. Four-week intervals were chosen because they allowed us to use Local Outlier Analysis to assess cluster and outlier patterns over our study period (this method requires at least 10-time slices), providing a more granular look at the temporal changes in the distribution of COVID-19 case rates across the city, while reducing time step temporal bias (bias that occurs from including additional dates based on the interval specified). (ArcGIS Pro, n.d.)

Our study ends on October 20, 2021, at the onset of the Omicron wave (fifth wave). As cases surged during this wave, government-sponsored laboratory testing for COVID-19 was limited to a selection of the population; thus, limiting the availability of comprehensive data on laboratory-confirmed COVID-19 cases for the entirety of the Omicron wave. (Public Health Ontario, 2021)

### Data Sources

Information on Toronto COVID-19 laboratory-confirmed case counts, location and timing of infection were identified through Ontario’s Public Health Case and Contact Management System (CCM) – a provincial surveillance database used to manage and report all laboratory-confirmed cases of COVID-19 cases and their contacts. (Ontario, 2022) Census tract-level demographic information (population counts, median total income, education attainment of residents, visible minorities, immigration and multigenerational housing) was identified through the 2021 Canadian Census. (Government of Canada, 2001) With the pandemic starting before the 2021 census, the CCM uses the 2016 census boundaries, which were the boundaries used to present our results. However, demographic information was adjusted to represent census information collected in the 2021 census to provide a more up-to-date representation of the city’s population.

### Statistical Analysis

To build a cohesive understanding of the dynamics of COVID-19 over space and time. we used three different applications of Moran’s I (Global Moran’s I, Local Moran’s I, and Local Outlier Analysis) because it is a widely used measure to identify spatial autocorrelation for health data and provides a cohesive approach to hotspot analysis in Toronto. Specifically, Global Moran’s I was used to evaluate whether COVID-19 case rates were spatially autocorrelated. (Anselin, 2020a; Cliff, 1973; Moran, 1950) Then, Local Moran’s I was used to identify clusters of the disease and Local Outlier Analysis was used to create a composite map that highlights the dynamic nature of COVID-19 in Toronto.

#### Global Moran’s I

Global Moran’s I was used to assess the presence of spatial autocorrelation in COVID-19 case rates at the city level. As a statistic grounded in Tobler’s First Law of Geography “*everything is related to everything else, but near things are more related than distant things*,” (Tobler, 1970, p. 236) we first defined a neighbourhood structure that relates each census tract to its neighbours. For this study, neighbours of each census tract were selected based on “queen” contiguity (neighbours share edges and corner boundaries) with neighbourhood relationships. (Anselin, 2022) This neighbourhood structure is then quantified through a weight matrix with row-standardized weights.

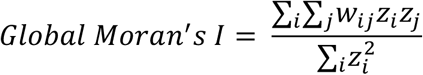

For Global Moran’s I, *w_ij_* is the spatial weight that relates census tract *i* to its neighbours *j*, *z_i_* (*z_i_* = *x_i_* − *x̄*) and *z_j_* (*z_j_* = *x_j_* − *x̄*) are the deviations of COVID-19 case rates at census tract i (*x_i_*) or neighbours j (*x_j_*) from the overall mean (*x̄*) COVID-19 case rate. (Anselin, 2020a)

Global Moran’s I value ranges from -1 to 1, with negative values closest to -1 indicating dispersion of case rates while positive values closest to +1 indicating clustering. A pseudo-*p-value* calculated through random permutations was used to assess whether the observed Global Moran’s I statistic is statistically significant. (Anselin, 2020a)

#### Local Moran’s I

Once spatial autocorrelation was established through Global Moran’s I statistic, Local Moran’s I was used to assess the clustering of COVID-19 case rates at a localized (census tract) scale for each of the four major waves of COVID-19. (Anselin, 1995) Local Moran’s I is a local version of Global Moran’s I that decomposes the impacts of overall geographical autocorrelation to localized spatial autocorrelation.

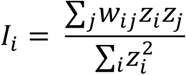

To calculate Local Moran’s I for census tract *i (I_i_)*, *w_ij_* is the spatial weight that relates census tract *i* to its neighbours *j, z_i_* (*z_i_* = *x_i_* − *x̄*) and *z_j_* (*z_j_* = *x_j_* − *x̄*) are the deviations of COVID-19 case rates at census tract *i* (*x_i_*) or neighbours *j* (*x_j_*) from the mean (*x̄*) of COVID-19 case rate.(Anselin, 2020b)

Local Moran’s I also rely on a neighbourhood structure to generate the spatial weight matrix. Neighbourhoods were also defined by “queen” contiguity with row standardized weights. A Local Moran’s I statistic was calculated for each census tract followed by random permutations to identify statistically significant clusters and outliers. Maps were generated to indicate areas of high-high (hotspots) and low-low (cold spot) clusters as well as outliers where COVID-19 case rate patterns did not match that of their surrounding area. (Fisman et al., 2021; Ontario, 2022)

#### Local Outlier Analysis

Local Outlier Analysis extends Local Moran’s I to identify clusters and outliers over both space and time, generating one map that shows the trends and changes of COVID-19 case rate concentration over space during the first two years of the pandemic. First, a space-time 3-D cube was generated to specify spatial and temporal relationships. The 3-D cube consists of space-time bins *(x, y, t)* representing COVID-19 case rates summed over a specified time period *(t)* for each census tract *(x,y).* (ArcGIS Pro, n.d.) Bins with the same time-step interval comprise a time slice, while bins with the same location (census tract) represent a time series.(ArcGIS Pro, n.d.)

For this method, we generated the 3-D space-time cube with each time slice representing a duration of 4 weeks (counted from the start date of January 20, 2020), resulting in a 3-D cube consisting of 572 bins (572 census tracts) x 23 -time steps (4-week intervals).

The specified 3-D cube was then used for the Local Outlier Analysis. Like the steps above, the neighbourhood structure for each bin was defined based on queen contiguity with random permutations used to identify statistically significant clusters and outliers over time. Each bin in a time slice was compared to the time slice preceding it to assess whether clustering patterns have changed over time. Results from this method provided a Local Moran’s I statistic for each bin as well as the number of hotspots, cold spots, and outliers over time. In addition, a map presentation of case rate patterns was created with a visual representation of the varying clustering characteristics over time (high-high, high-low outlier, low-high outlier, low-low cluster, and multiple types – indicating different statistically significant patterns over time) (<u>See</u> Table 4).

**Table 4:** Local Outlier Analysis provides a succinct depiction of changing clustering patterns in a location over a specific duration of time. Census tracts that have continuously been COVID-19 hotspots or cold spots from January 2020 – October 2021 are identified as light pink or light blue, respectively. Census tracts that have continuously been outliers compared to surrounding census tracts are identified as bright red for high-low outliers and bright blue for low high outliers. Census tracts that have had changing clustering patterns over the study period are identified as purple (ex. one time slice the census tract was a hot spot and became a cold spot in another time slice). Census tracts without any statistically significant clustering patterns throughout the study period were identified as white.

| **Table 4: Local Outlier Analysis Clustering Pattern** (ArcGIS Pro, n.d.) |  |  |
| --- | --- | --- |
| Clustering pattern |  | Definition |
|  | Never Significant | A location where there has never been a statistically significant clustering pattern |
|  | Only High-High Cluster | A location where the only statistically significant clustering pattern throughout time has been high-high clusters. |
|  | Only High-Low Outlier | A location where the only statistically significant clustering pattern throughout time has been high-low outliers. |
|  | Only Low-High Outlier | A location where the only statistically significant clustering pattern throughout time has been low-high outliers |
|  | Only Low-Low Cluster | A location where the only statistically significant clustering pattern throughout time has been low-low clusters. |
|  | Multiple Types | A location where the statistically significant clustering pattern throughout time changes |

#### Comparative Map Analysis

In addition to maps generated from Moran’s I applications, individual maps were generated for social demographic characteristics of interest (Table 1) to compare with hot spot maps and explore potential relationships.

#### Sensitivity Analysis

To ensure the robustness of our results given different spatial structures we revisited our main analysis with a few modifications. Our first modification was to the neighbourhood structure, which plays a crucial role in the results produced in spatial analysis. Instead of queen contiguity, we repeated our analysis with two fixed distances, identified using the ArcGIS Incremental Spatial Autocorrelation tool to find the appropriate distance thresholds for hot spot analyses. (ArcGIS Pro, n.d.) Two distances were identified: a distance to ensure each census tract had at least one neighbour as well as the distance that ensures the most pronounced spatial autocorrelation. (ArcGIS Pro, n.d.) To address the potential issue of modifiable areal unit problem (MAUP), where results vary based on the areal unit scale, our second modification was to use Toronto neighbourhoods instead of census tracts. Toronto Neighbourhood boundaries combine 2-5 census tracts of similar socioeconomic status to help Toronto Municipality plan and implement resources at a meaningful governmental scale, making it an appropriate areal unit to scale our analysis beyond census tracts. (City of Toronto, 2022b)

Beyond the neighbourhood structure, we also stratified our analysis by biological risk factors for COVID-19 like age (older than 65 and less than 65) and sex assigned at birth (male and female) to identify any differences that might have been missed. (Booth et al., 2021)

All analyses were completed in ArcGIS Pro (version 2.9.3) and R statistical software (version 4.1.3)

### Ethics Approval

This study has been approved by the University of Toronto Health Sciences Research Ethics Board (protocol number 44167).

## RESULTS

### Primary Analysis

Global Moran’s I was used to assess the presence of spatial autocorrelation across the five major waves in the first two years of the pandemic. We identified statistically significant positive spatial autocorrelation across all four major waves, indicating spatial clustering of cases. The resulting Global Moran’s I value for each wave can be found in Table 5.

**Table 5:** Global Moran’s I Index values for each COVID-19 wave, indicating the presence of spatial autocorrelation. Positive index values indicate spatial clustering of COVID-19 cases with values closer to 1 indicating higher levels of clustering

| **Table 5: Global Moran's I Index for four COVID-19 waves in Toronto from January 2020 – October 2021** |  |  |  |
| --- | --- | --- | --- |
| Study Period/Waves | Global Moran's I Index | Z Score | p-value |
| Wave 1 – Period 1 | 0.430 | 18.40 | <0.001 |
| Wave 2 – Period 2 | 0.544 | 23.11 | <0.001 |
| Wave 3 – Period 3 | 0.496 | 21.04 | <0.001 |
| Wave 4 – Period 4 | 0.193 | 8.369 | <0.001 |

Local Moran’s I was used to identify clusters of COVID-19 case rates at the census tract level (<u>See</u> Figure 2). The first three major waves showed similar patterns. Hotspots were concentrated in the northwestern part of Toronto, and later on in the northeastern corner of the city in the second and third wave. Meanwhile, COVID-19 cold spots were identified lining midtown from north to south with a few cold spots situated in the southwestern part of the city. The pattern of low-high outliers remained relatively consistent across the first three waves while high-low outliers varied slightly in pattern over the first three waves.

**Figure 2:**
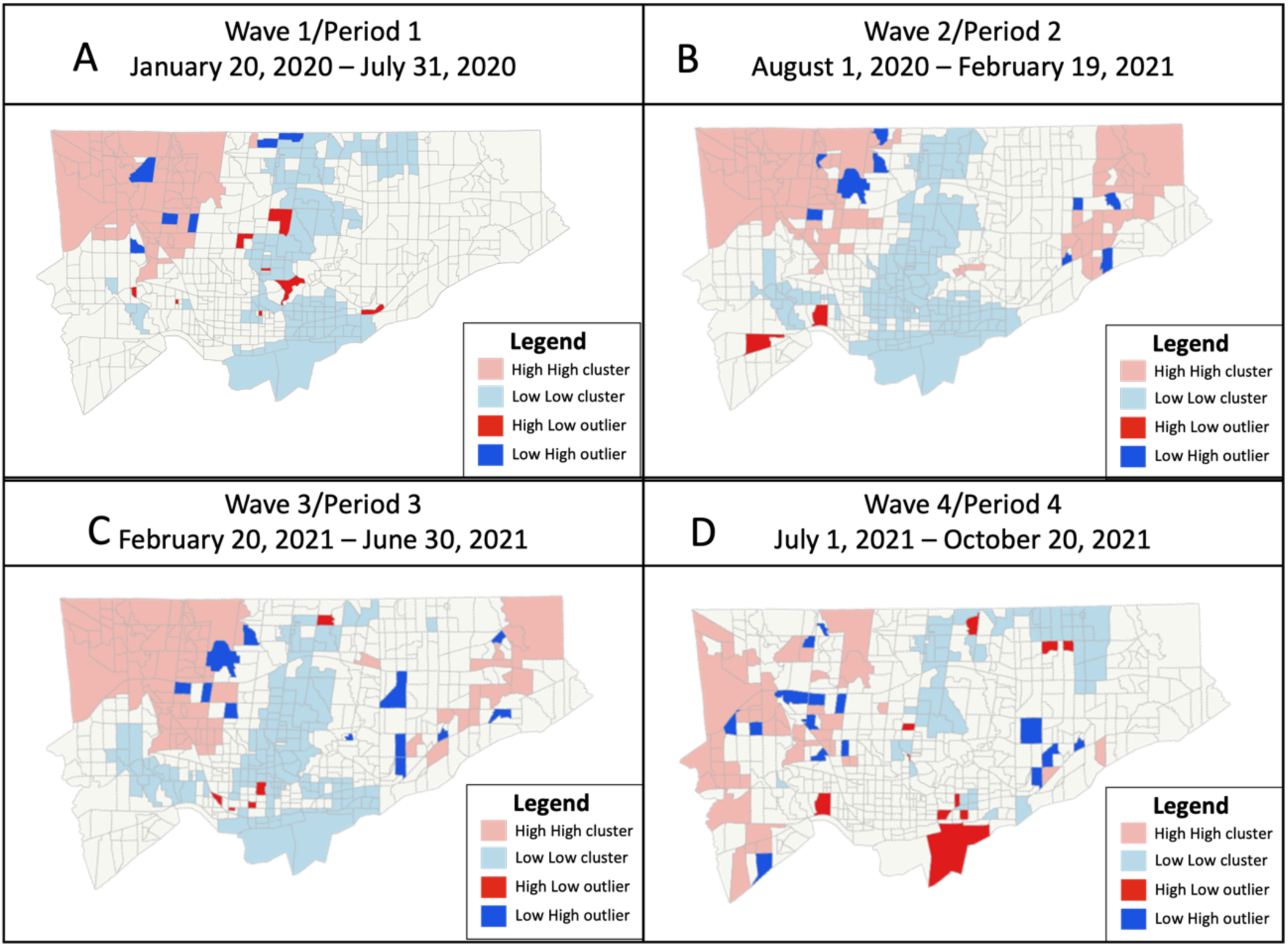
COVID-19 case rate (per 100,000) clustering patterns across Toronto for each major waves using Local Moran’s I.

By the fourth wave (Figure 2D), hotspots had shifted south and were found along the western border of the city. More cold spots were now found in the northern part of the city. There were also more outliers in this wave.

Local Outlier Analysis allowed us to create a composite map that highlighted the dynamic nature of COVID-19 over space and time (Figure 3A). Over the course of the pandemic, on average, cold spots (coloured blue) remained midtown with an extension to the southeastern part of the city. Upon further inspection, most of the census tracts in these cold spot zones that were identified as having multiple clustering patterns over time were largely cold spots for most of the study period (Figure 3B). Despite the northern corners of the city showing the most variable clustering pattern over space and time, most of the time slices indicated hot spots (Figure 3C-3D).

**Figure 3:**
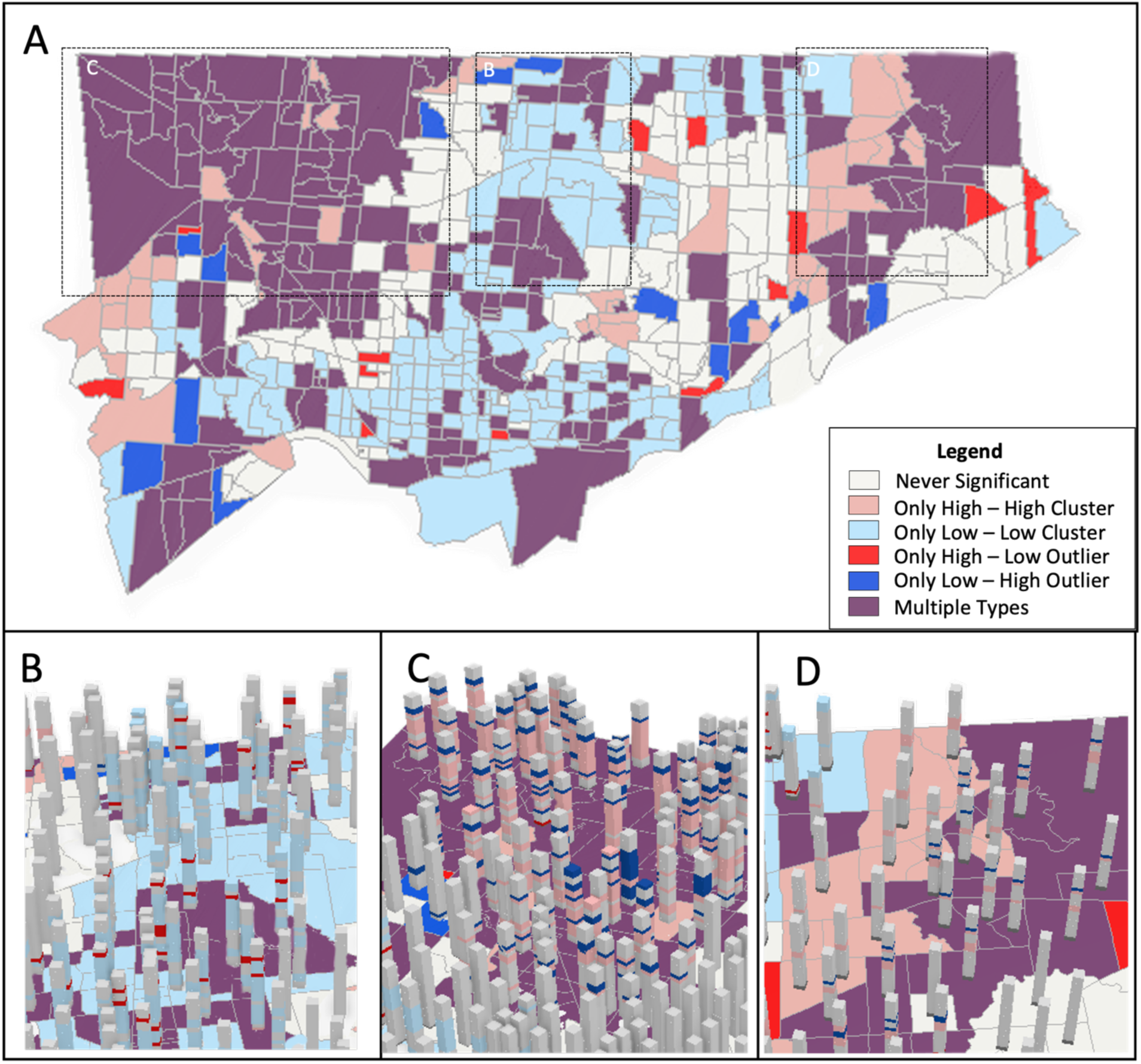
A. COVID-19 case rate (per 100,000) clustering patterns from January 2020 – October 2021 analyzed using Local Outlier Analysis. B-D. Time slices in sections of the city with multiple types of clustering and outlier patterns.

Comparing hotspot maps to social demographic maps (Figure 4A-E), general trends were apparent. Hotspots were found in areas with a larger proportion of residents who identified as visible minorities and immigrants, were less educated and lived in multigenerational homes.

**Figure 4:**
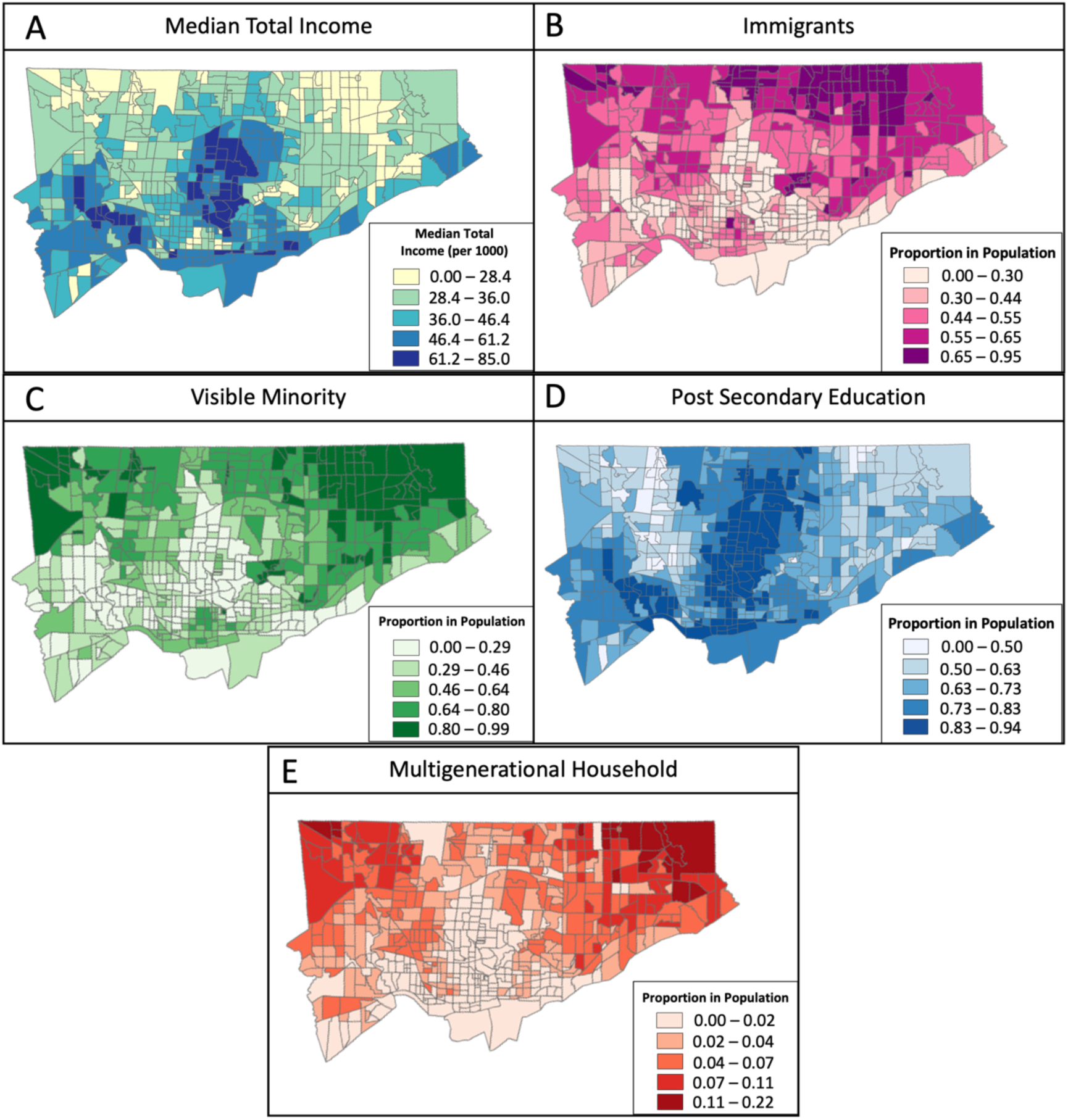
Spatial distribution of sociodemographic and socioeconomic characteristics in Toronto based on the 2021 Canadian Census. A. Distribution of Median Total Income per individual. B. Distribution of Immigrants C. Distribution of Visible Minorities. D. Distribution of residents with a Post Secondary education. E. Distribution of multigenerational households in Toronto.

### Sensitivity analysis

We repeated our analysis with a few modifications to assess whether the trends identified were consistent irrespective of how the neighbourhood was defined or according to various key demographic information. Regardless of how we defined our neighbourhood structure (fixed distance or neighbourhood level) or how we stratified our population based on biological risk factors, our results were consistent. When we changed the neighbourhood structure to be defined based on two fixed distances (2,277m and 8,854m) instead of queen contiguity, patterns for Local Moran’s I (Appendix Figure 1-4) and Local Outlier Analysis (Appendix Figure 5), were also similar to our primary analysis; however, the shorter distance, and more so the longer distance, were not as specific, with most census tracts indicating either cold or hot spots. For the Toronto neighbourhood-level analysis, patterns were also consistent across the four waves, with slight variation. The major difference was that Toronto neighbourhood-level analysis did not capture any statistically significant hotspots in the northeast corner of the city during the first three waves. The slight variation in results was not unexpected given the neighbourhood level averages trends over a larger distance compared to the census tract level.

We also stratified our analysis by COVID-19 two individual characteristics, sex assigned at birth (male and female) and by age category (less than 65 and over 65) where we identified consistent positive spatial autocorrelation consistent to our primary analysis, irrespective of age and sex. For Local Moran’s I and Local Outlier Analysis, all maps generated were also consistent to our primary analysis with patterns of hot spots, cold spots and outliers consistent irrespective of age and sex. Maps generated with only adults older than 65 years had fewer statistically significant clustering patterns because of the fewer individuals in this age group, possibly because LTC residents, who tend to be in this age group, were not included in the analysis.

## DISCUSSION

Our hot spot analysis described the dynamic nature of COVID-19 case rates across the city of Toronto from January 2020 to October 2021. In the first three waves of the pandemic, COVID-19 case rates were concentrated in the northern corners of the city. By the end of the fourth wave, the COVID-19 landscape had changed, with hotspots shifting southward. Comparing spatial patterns to social demographic factors, the demographic makeup of a census tract might have had some relationship to the intensity of COVID-19 rates identified.

Comparing our results to current literature, temporal changes in spatial distributions of COVID-19 have been previously addressed in studies in the US, demonstrating both persistent and evolving patterns of COVID-19 cases and outcomes over space and time connected to neighbourhood-level social and demographic characteristics. (Neelon et al., 2021; Park et al., 2021; Tsou et al., 2023) Spatial studies in Toronto also identified similar spatial patterns with persistent hotspots in the northwestern corner of the city and cold spots lining the centre of the city for most of 2020 and the first half of 2021. (Forsyth et al., 2023; Mishra et al., 2022; Nazia et al., 2022; Vaz, 2021) Higher COVID-19 case rates were also identified to be localized in areas of Toronto with more multigenerational households and a higher concentration of lower-income, immigrant, visible minority and less educated residents. (Abdi et al., 2021; McKenzie, 2020; Nazia et al., 2022; Mishra et al., 2022; Forsyth et al., 2023; Wang et al., 2023) Our study adds to the current literature in a few ways. By focusing on each wave, we were able to identify additional hotspots in the northeastern part of the city and provide individual snapshots of how different each wave was spatially. With a composite hot spot map using Local Outlier Analysis, a tool that has not been used in current literature, we were also able to provide a succinct depiction of how COVID-19 case concentrations changed over space and time, while comparing spatial patterns to that of sociodemographic and socioeconomic characteristics of the city.

Comparing spatial patterns to social demographic factors, the demographic makeup of a census tract might have had some relationship to the concentration of COVID-19 identified. Higher COVID-19 case rate concentration in areas of lower income, higher immigrant, higher visible minorities and less educated populations with more multigenerational households are corroborated in other studies of the pandemic experience in Toronto(Abdi et al., 2021; McKenzie, 2020; Nazia et al., 2022; Mishra et al., 2022; Forsyth et al., 2023; Wang et al., 2023) Sociodemographic and socioeconomic differences dictate the different physical/built environments residents of Toronto find themselves in, which may interact to impact the risks of acquiring COVID-19 and experiencing adverse outcomes if infected. For instance, low-income communities, which also often have higher concentrations of immigrants, tend to have larger household sizes and a higher proportion of multigenerational housing, which may limit personal space for social distancing, increase the risk of spreading respiratory infectious diseases within households, as well as facilitate spread of infection between age groups (e.g., from school children to their grandparents). (Elliot and Leon, 2020; Forsyth et al., 2023; Hassen, 2021; Nazia et al., 2022; O’Neill et al., 2022) Furthermore, these communities also tend to have a higher concentration of residents that provide essential services – working as personal support workers, in food supply, at warehouses, and in retail—often with limited access to personal protective equipment, lack of paid sick leave and fewer options to work from home or “shelter-in-place”. (Forsyth et al., 2023; Hassen, 2021; Mishra et al., 2022; Nazia et al., 2022; O’Neill et al., 2022; Rao et al., 2021) These *essential workers* often rely on public transit to and from work and thus are potentially exposed to SARS-CoV-2, not only at home but at work and in transit. (Forsyth et al., 2023; Hassen, 2021) Public health initiatives introduced in the first year of the pandemic to potentially reduce the risk of COVID-19 might not have adequately reduced the burden of COVID-19 in more marginalized Torontonians throughout the waves.(Ma et al., 2022)

Focusing on the fourth wave, popularly described as the Delta wave, the changing spatial distribution of COVID-19 case rates towards the end of 2021 was mainly driven by a more transmissible and infectious variant – B.1.617, during the summer of 2021, along with relaxed public health restrictions and pandemic fatigue that saw fewer behavioural changes even as cases were rising again.(Anthes, 2021; Lao, 2021; Pelley, 2021) Spatially, COVID-19 case rates were still concentrated in more marginalized populations but also included hotspots in census tracts that had fewer minorities, higher median income, fewer multigenerational homes and more educated residents. Studies have postulated that strict public health guidelines implemented earlier in the pandemic might disproportionally kept the more affluent, better-educated and white individuals at home, potentially reducing their risk of COVID-19. (Huang et al., 2022, 2021; Iio et al., 2021) Once public health guidelines were relaxed at the onset of the Delta wave, overall population mobility increased (Alam, 2021; Dai et al., 2023; Pelley, 2021), which might have also explained the addition of hotspots in census tracts with a more privileged population.

Aside from the general trends identified, one unique pattern was identified in our maps that did not particularly conform to the overall trends highlighted in the literature. There were several census tracts located in the northwestern part of the city that had on average, lower COVID-19 case rates, especially waves 1 and 4. As noted above, these census tracts have a higher proportion of immigrants and low-income residents, but after further investigation, these areas of the city also have a higher proportion of residents identifying as East Asian.(Government of Canada, 2022) Toronto Public Health identified a similar trend of lower SAR-CoV-2 infection rates while assessing case count differences among major visible minority groups in the City of Toronto. (City of Toronto, 2020) Given the increase of anti-Asian racism affecting the East Asian community during the pandemic, it was notable that COVID-19 case rates appeared disproportionately lower amongst this demographic. Studies focusing on the Chinese Canadian community in the Greater Toronto Area have highlighted cultural factors such as collectivism (placing the community above the individual), increased information-seeking behaviour through culturally specific social media platforms, the cultural symbolism of mask-wearing (a useful multifunctional tool rather than a sign of contagion), social ties in China that provided early alert of the severity of COVID-19, and prior experience with SARS in 2003, as some of the unique cultural factors that might have promoted early vigilance against COVID-19 and overall lower COVID-19 cases. (Lee et al., 2021; Mamuji et al., 2021) However, more culturally sensitive research looking at various East Asian groups in Toronto is needed to further elucidate this trend.

Our study has many strengths in providing snapshots over time of the COVID-19 pandemic in Toronto during the first two years; however, it has some limitations. First, our study is descriptive and focused on exploring potential indicators that might have dictated the spatial and temporal distribution of COVID-19 cases in Toronto. Patterns do not mean concrete evidence for correlation, and we did not formally assess associations or correlations. Rather, our results will inform future more complex spatial and temporal Bayesian analysis of COVID-19 case rate distribution in Toronto.

Second, COVID-19 testing might have impacted our ability to truly assess the extent of the pandemic experience for all Torontonians. As the Delta variant spread quickly in the summer of 2021 to dominate the fourth wave, and more aggressively, Omicron by December 2021, provincial testing was beyond capacity, resulting in severe restrictions in laboratory SARS-CoV-2 testing and increasing case backlogs, which resulted in us restricting our analysis to the end of October 2021. (White, 2021) Unfortunately, our current case rate data was not adjusted for testing but the next step for this project will be to consider testing variability across the city to better adjust for varying COVID-19 case counts.

Third, like many spatial studies, our study is subject to MAUP. Although we tried to address this problem with additional analysis at the neighbourhood level, we were limited to two areal units (census tract and neighbourhood level), given the collection of COVID-19 case data. Since the COVID-19 pandemic was underway a full year before the 2021 census, the CCM used the 2016 census tract demarcation to connect cases to census tracts, which was easily adjustable to account for population changes identified in the 2021 census. This was a similar situation for the Toronto neighbourhood demarcation, even as the city transitioned to a newer neighbourhood structure in 2022. However, making similar adjustments was challenging for area units such as Forward Sortation Area (FSA), another important geographical unit of analysis, that is updated more frequently between census cycles and harder to adjust population counts without full residential addresses. Nevertheless, assessing COVID-19 case rate trends across two areal units allowed us to assess the robustness of our results and how they could change over different areal units.

## CONCLUSION

Our study provides a necessary in-depth look at the COVID-19 epidemic in Toronto over space and time, with an emphasis on the dynamic nature of the pandemic. Although many now believe that the pandemic is largely over, the COVID-19 pandemic has shone a spotlight on the glaring structural health inequities in Toronto. There is still more to learn about the nuanced experience of Torontonians as the conversation of its irreversible impacts on society and population health is gaining more interest, especially as long-COVID is of public health concern. Investigating the early years of the pandemic with a more detailed and nuanced look helps us add more detail to the growing literature focused on understanding the connection between social inequities and increased COVID-19 risk for the marginalized members of the city.

## Data Availability

Data cannot be shared publicly due to data sharing agreements and legal restrictions. Census Data is publically available through Statistics Canada.

## APPENDIX

**Appendix Figure 1:**
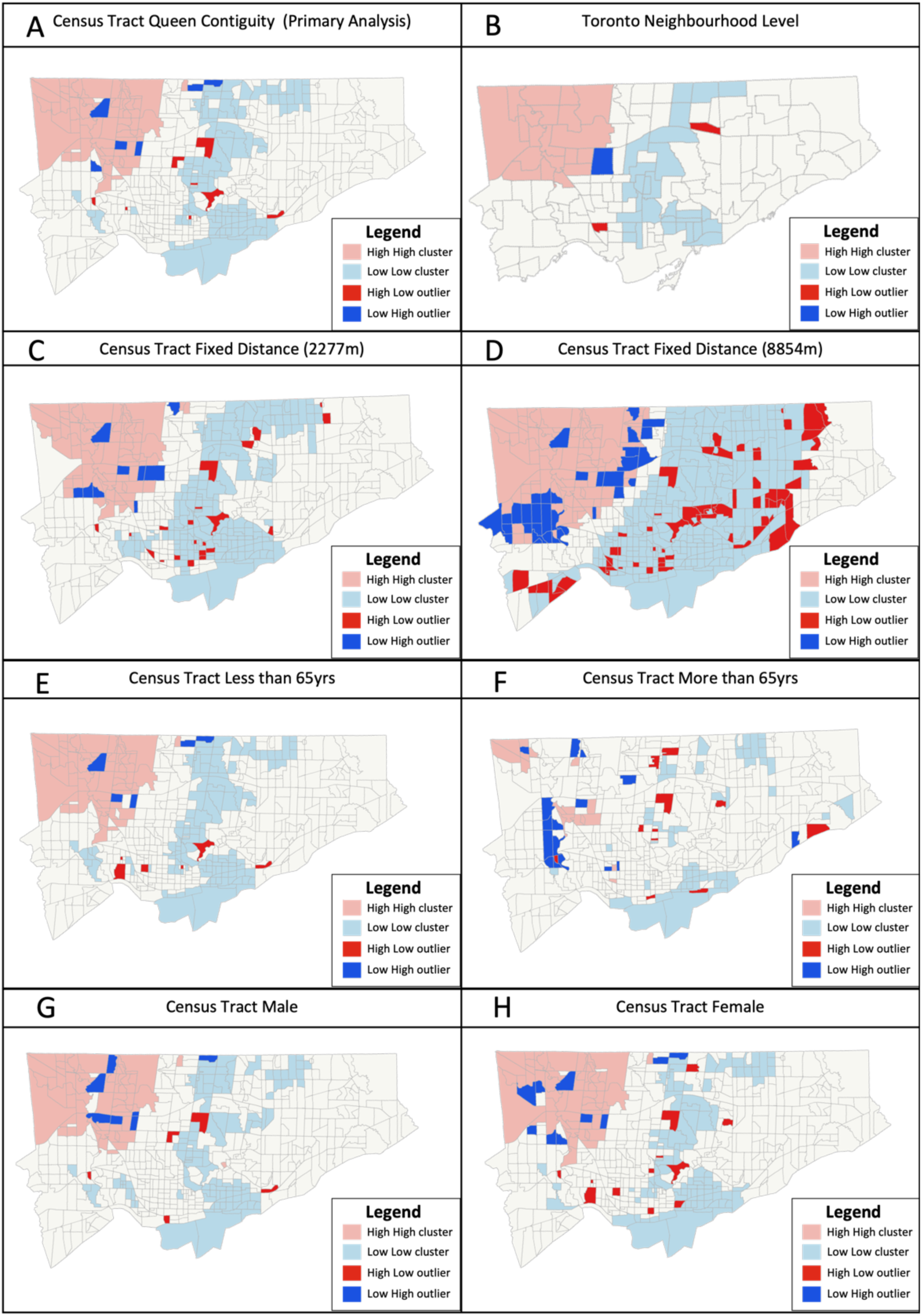
COVID-19 case rate (per 100, 000) clustering pattern across Toronto for Wave 1 with varying population characteristics (age -- E&F and sex – G&H) and neighbourhood structure (fixed distance – C &D and neighbourhood level B) adjustments compared to map generated during primary analysis (A).

**Appendix Figure 2:**
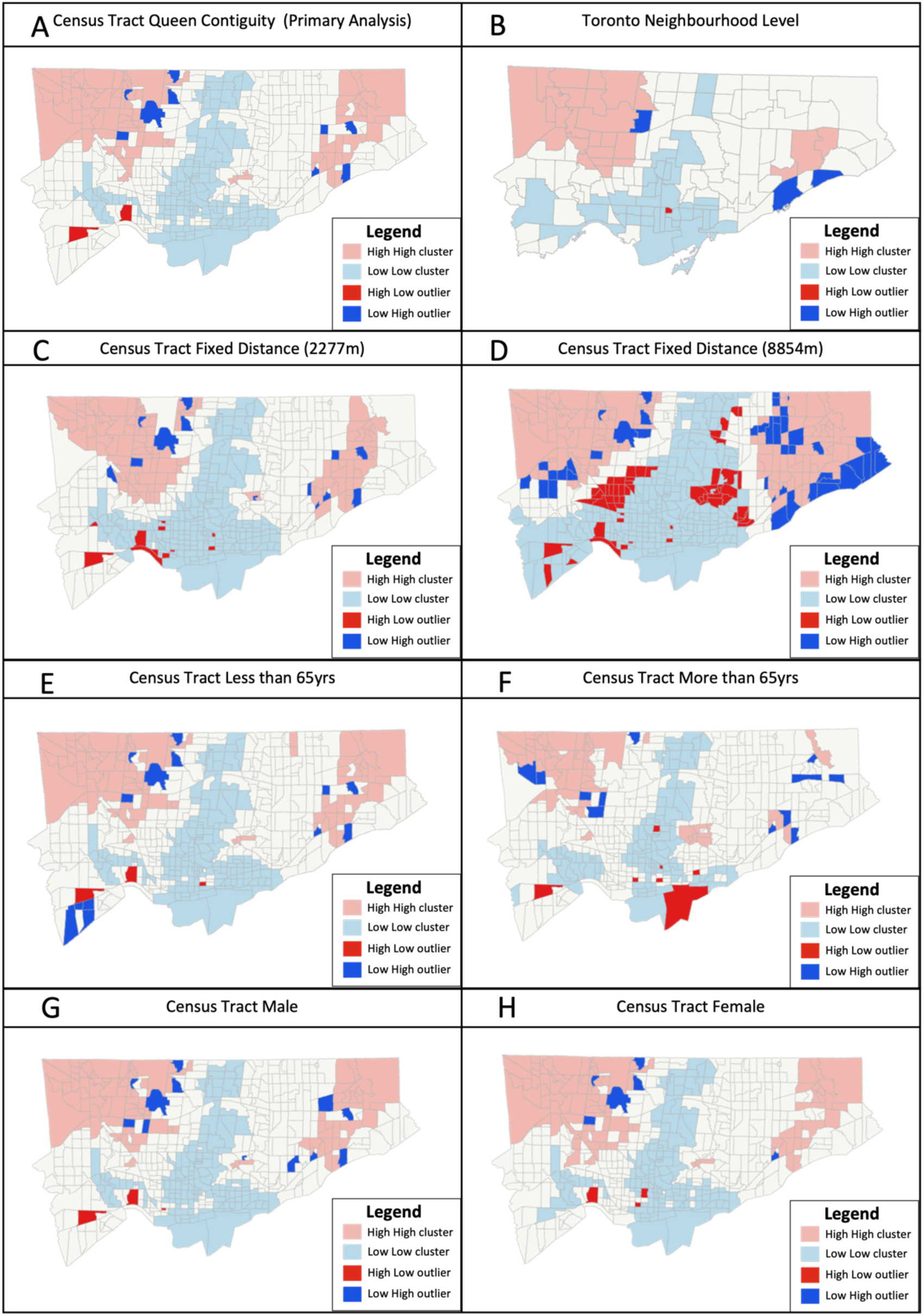
COVID-19 case rate (per 100, 000) clustering pattern across Toronto for Wave 2 with varying population characteristics (age – E&F and sex – G&H) and neighbourhood structure (fixed distance – C &D and neighbourhood level – B) adjustments compared to map generated during primary analysis (A).

**Appendix Figure 3:**
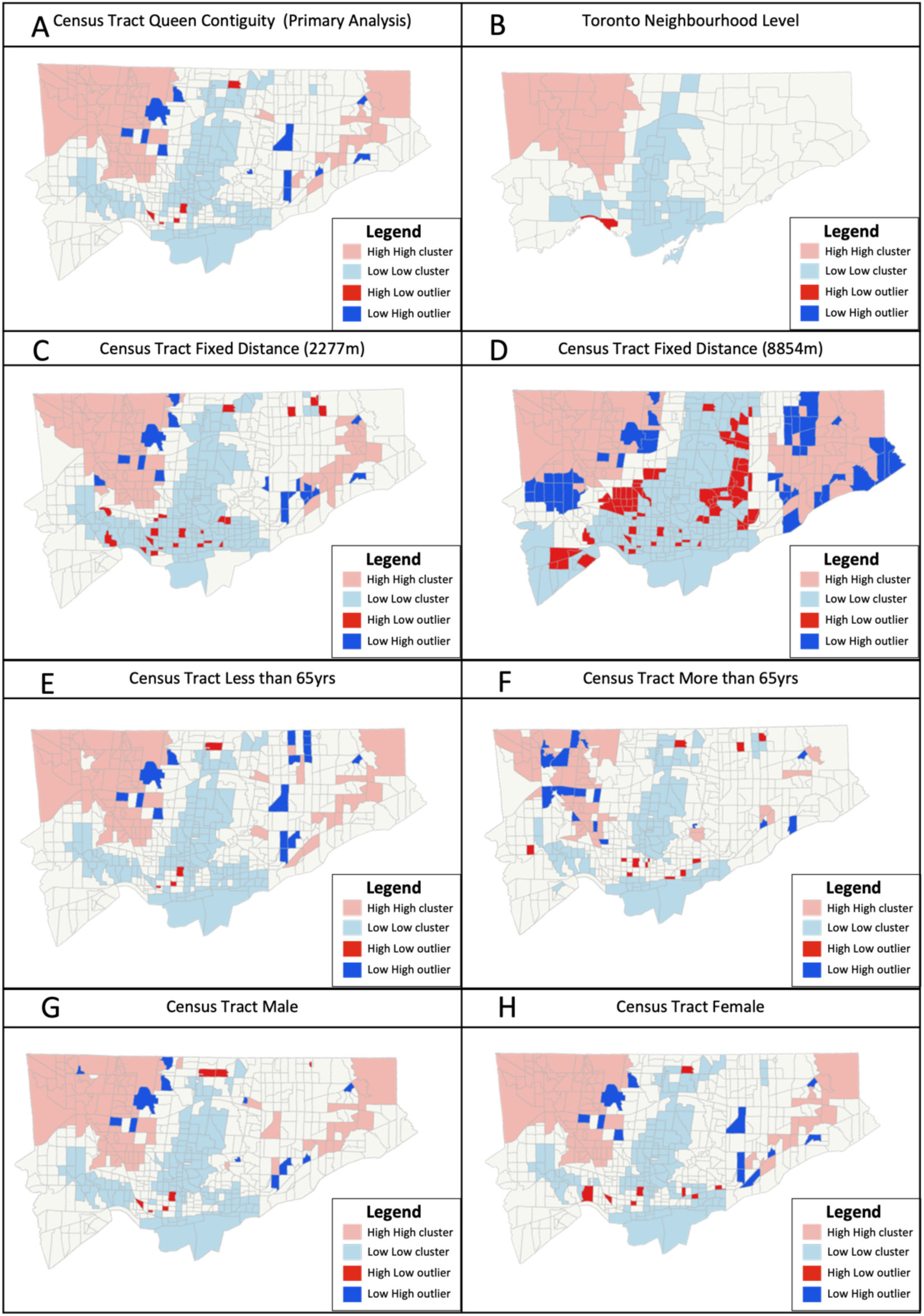
COVID-19 case rate (per 100, 000) clustering pattern across Toronto for Wave 3 with varying population characteristics (age – E&F and sex – G&H) and neighbourhood structure (fixed distance – C &D and neighbourhood level – B) adjustments compared to maps generated during primary analysis (A).

**Appendix Figure 4:**
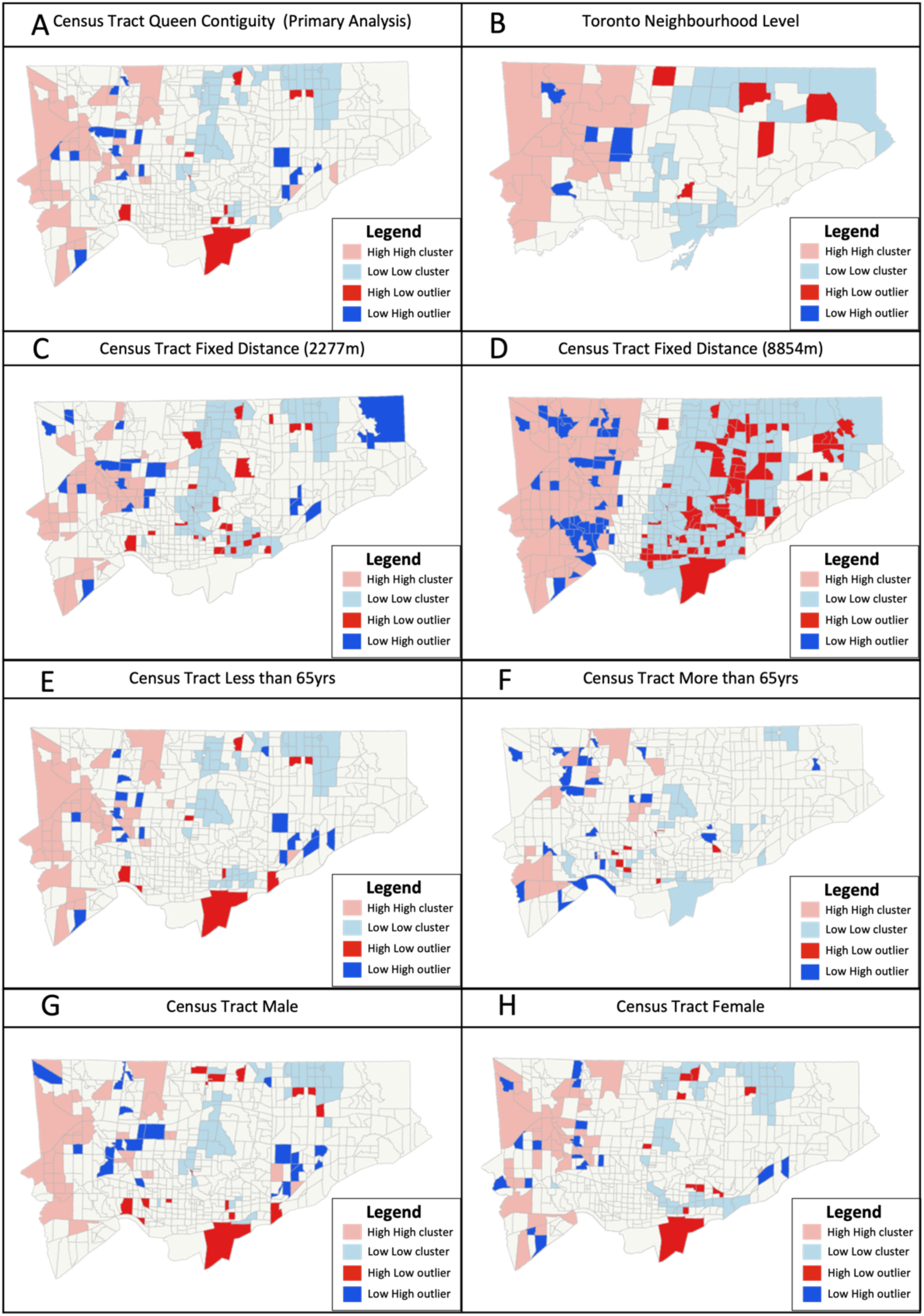
COVID-19 case rate (per 100, 000) clustering pattern across Toronto for Wave 4 with varying population characteristics (age – E&F and sex – G&H) and neighbourhood structure (fixed distance – C &D and neighbourhood level B) adjustments compared to maps generated during primary analysis (A).

**Appendix Figure 5:**
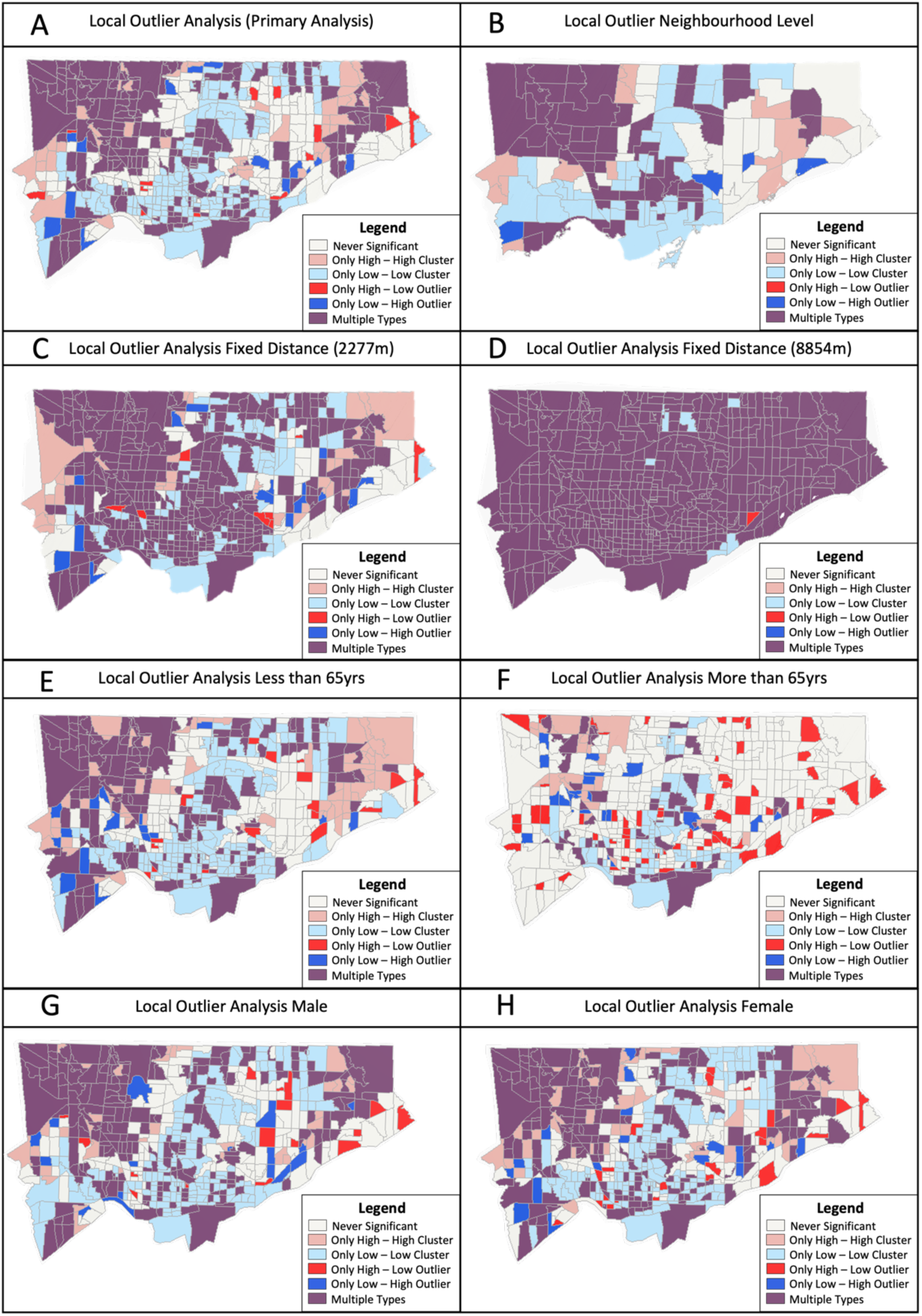
COVID-19 case rate (per 100,000) clustering patterns from January 2020 - October 2021 analyzed using Local Outlier Analysis with varying adjustments focusing on population (age – E&F and sex – G&H), neighbourhood structure (fixed distance – C &D and neighbourhood level – B) as compared to primary analysis (A).

